# The Case for Altruism in Institutional Diagnostic Testing

**DOI:** 10.1101/2021.03.16.21253669

**Authors:** Ivan Specht, Kian Sani, Yolanda Botti-Lodovico, Michael Hughes, Kristin Heumann, Amy Bronson, John Marshall, Emily Baron, Eric Parrie, Olivia Glennon, Ben Fry, Andrés Colubri, Pardis C. Sabeti

## Abstract

Amid COVID-19, many institutions deployed vast resources to test their members regularly for safe reopening. This self-focused approach, however, not only overlooks surrounding communities but also remains blind to community transmission that could breach the institution. To test the relative merits of a more altruistic strategy, we built an epidemiological model that assesses the differential impact on case counts when institutions instead allocate a proportion of their tests to members’ close contacts in the larger community. We found that testing outside the institution benefits the institution in all plausible circumstances, with the optimal proportion of tests to use externally landing at 45% under baseline model parameters. Our results were robust to local prevalence, secondary attack rate, testing capacity, and contact reporting level, yielding a range of optimal community testing proportions from 18% to 58%. The model performed best under the assumption that community contacts are known to the institution; however, it still demonstrated a significant benefit even without complete knowledge of the contact network.

## 1 Introduction

During any societal crisis, altruism has the potential to both satisfy moral duty and maximize “utility,” leading to the best possible outcome for the greatest number of people [24]. It gains newfound urgency and utility during a pandemic, when important decisions must be made around allocating scarce resources, such as tests, therapies, and vaccines. In these instances more than ever, our own interests––our health, safety, and well-being––become highly interdependent on those of others. Specifically for communicable diseases, testing is patently a public good because a positive result can reduce others’ exposure and suffering by guiding isolation and quarantine practices.

Considerations of altruism and its efficacy have resurfaced in various COVID-19 response plans worldwide. As the disease began to spread in the U.S., it forced schools and businesses to cease in-person operations to mitigate its spread. To reopen, many of these institutions rushed to test their own members, enacting several-times-per-week or even daily testing protocols in hopes of preventing outbreaks [15]. Countless institutions spent millions on internal testing programs. Some universities, for instance, spent upwards of $1-2 million per week to test students and staff, often using clinical-grade diagnostics [3, 20]. Meanwhile, communities surrounding these institutions continued to struggle with ongoing clinical testing shortages and long delays for results. Even for institutional testing programs that considered supporting community testing, legal and regulatory barriers served as an additional deterrent from doing so.

To turn inward is a common and understandable approach during any crisis, but these expensive self-focused testing programs still left institutions blind to community cases that could potentially enter and spread like wildfire. For example, the NFL spent $100 million in total throughout the 2020-21 season on nearly one million tests for around 7,500 institutional members, administered daily [19]. Yet, the League still experienced several outbreaks [22]. They were not alone; outbreaks occurred within many similar testing programs, as the world witnessed prominently at the White House in Fall 2020.

We are now faced with the question of whether the confined use of significant resources to enable high-frequency testing within individual institutions alone is the most appropriate or effective way to contain a virus. We hypothesized that if institutions test altruistically––that is, designate a substantial portion of their testing capacity outside the institutions––it would not only be good for their communities, but also for them. That is, there would be lower case counts in these institutions themselves had their programs incorporated the testing of close contacts of its members into its testing strategy, thereby detecting potential COVID-19 encroachment.

This paper seeks to ascertain whether a self-focused or an altruistic testing approach is a more effective mitigation strategy. We provide a simple yet plausible epidemiological model to answer this question, comparing results under varying local community prevalence levels, social mitigation efforts, testing capacity, contact tracing adoption, and other parameters. We then discuss the significant real-world implications of our findings concerning how institutions might better allocate their available testing capacity

## 2 Epidemiological Model

To test our central claim, we built an agent-based epidemiological model of a hypothetical institution such as a workplace, school, or similar organization, accounting for interactions between within the institution as well as between the institution and its surrounding community. For a full, mathematically rigorous methods section, see Appendix A; here, we provide an intuitive, high-level description. We modeled two distinct groups: (1) institution members and (2) all of their first-degree close contacts outside the institution (hereafter referred to as the ‘periphery’). We assumed that the periphery remains unchanged throughout the simulation. We then assessed the effect of redistributing some of the institution’s testing capacity to the periphery, assuming for simplicity that diagnostic testing in the community was negligibly rare before the institution’s intervention. The model provided critical insight into the optimal proportion of tests to redistribute, given several baseline assumptions about viral dynamics, prevalence, and more. Moreover, we assumed no knowledge of the institution and the periphery beyond what institutional administrators/health officers might reasonably gather, such as the number of individuals, their frequent contacts, and the number of tests conducted. As such, our model gives a general framework by which institutions may assess possible testing protocols’ effectiveness.

Modeling viral propagation between an institution and its periphery requires detailed information on how the agents involved interact with each other. A typical means to capture this information is a simple undirected graph, in which two nodes (i.e., people) share an edge if and only if those two people interact during the modeled period [1]. In a real-world context, we might construct this graph by, for example, surveying members of the institution and its surrounding community about their social interactions. For our model, however, we assumed knowledge only of the mean and variance in contact numbers inside and outside the institution, which is likely more feasible to estimate in most contexts.

We proceeded by modeling *N* agents who interact according to a random graph, in which node degrees follow an overdispersed Negative Binomial distribution fit to the observed mean and variance (see Appendix A for the random graph generation algorithm) [12]. To account for institution-periphery interactions, we assigned each agent a number of regular contacts made outside the institution, drawn from a different Negative Binomial distribution. See Figure 1a for an example of a contact network generated via this method. This contact network may or may not be fully known to the institution; we included a model parameter that captures the extent to which agents report their contacts.

**Figure 1:**
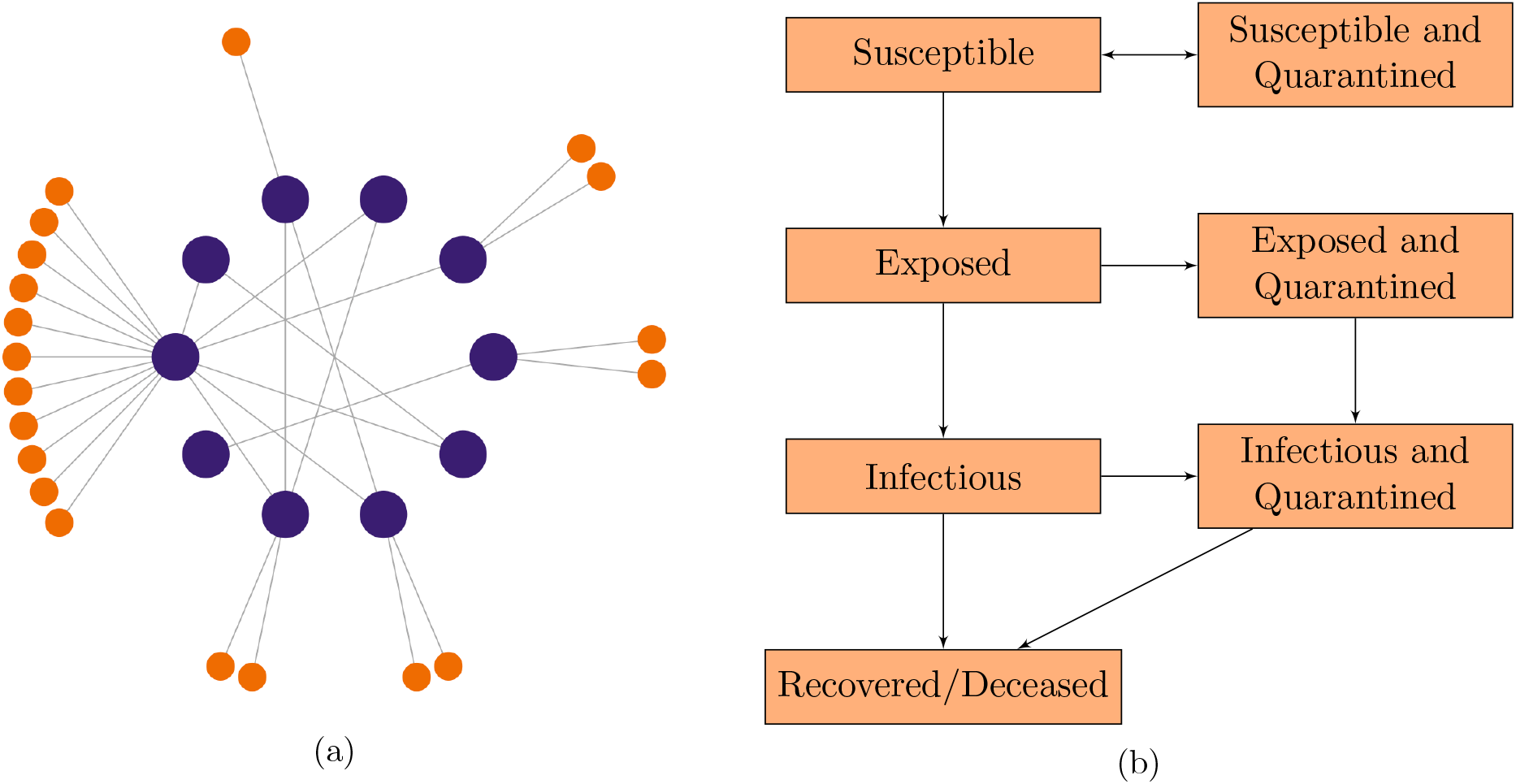
(a) Example of a contact network representing members of the institution (large, purple nodes) and their contacts in the periphery (small, orange nodes). Here we have 10 institution members who make an average of 2 contacts within the institution and 2 contacts outside the institution (variance = 3 for both distributions). (b) Flowchart of compartments and possible state transitions.

Having established a graphical contact network, we experimented with different testing strategies to simulate the spread of COVID-19. As a first step, we assumed that the periphery exhibits an epidemiological steady-state, computed based on the proportion of tests distributed there. By steady-state, we mean that the probability of an individual in the periphery being in a certain epidemiological state remains constant over the course of the simulation (see Appendix C for a more detailed treatment of this assumption). Within the institution, by contrast, we set the initial infection rate low in comparison to the periphery, reflective of the fact that many institutions returning to in-person activities have rigorous testing/quarantining protocols. For these agents, we modeled viral states probabilistically via an adapted *N*-intertwined mean-field approximation (NIMFA). In its original form, the NIMFA states that at a given time, the rate of transmission from agent *j* to adjacent agent *i* is proportional to the product of the marginal probabilities that *i* is susceptible and that *j* is infectious [17, 18]. This approach offers the flexibility to encompass a wide range of epidemiological compartments while capturing the granularity of graphical contact networks [13, 21, 25].

We then extended NIMFA to include testing-based interventions. First, we allowed the outgoing transmission rate to vary between agents as a partial means of modeling overdispersion (with the rest coming from the node degree distribution). To account for quarantine compartments, we set the time-dependent rate at which agent *j* enters quarantine proportional to the product of the marginal probabilities that *j* is infectious and that *j* receives a test. In turn, this latter probability depended on the test distribution strategy, the probability that *j* had not previously tested positive, and the number of adjacent nodes in *j*’s contact network. Finally, accounting for the fact that COVID-19 cases exhibit an exposed (but not yet infectious) stage, we arrived at detailed compartmental model, in which agents transition between epidemiological states as depicted in Figure 1b.

## 3 Results

We first applied our model to a mid-sized university (*N* = 10,000), using real data we gathered at Colorado Mesa University (CMU). CMU established a testing program in summer 2020 initially focused on university students and staff, and began supporting testing in the greater Mesa County community later that year [4]. Contact tracers determined that the mean and variance of the number of close contacts within the institution were 2.3 and 2.4, respectively, and outside the institution were 0.2 and 1.9, respectively. They also found that the prevalence on campus at the beginning of the Spring 2020 semester was approximately 1%, and that they planned to conduct about 0.12 diagnostic tests per day per person. Supplementing our own data collection with that of the local public health authority, we compiled a complete set of parameters specific to CMU and ran the model accordingly (see Appendix D).

Our initial model results based on the CMU parameters provided strong evidence in support of an altruistic testing strategy. We observed that the projected number of cases 40 days after the beginning of the modeled period was lowest when CMU deployed 45% of its tests to the periphery (see Figure 2). This strategy reduced the institutional case count by 25% as compared to a self-focused testing strategy (i.e. 0% peripheral testing). However, our data from CMU––which informed our baseline parameters––were likely subject to several biases. While CMU administrators attested that they believed the data to represent the student body fairly well, students who contracted the virus (as every individual in our dataset did) were likely to have higher degrees of social interaction than those who did not, leading to a positive bias. On the other hand, CMU informed us that certain close contacts were likely not reported or otherwise not included in some cases, introducing a negative bias.

**Figure 2:**
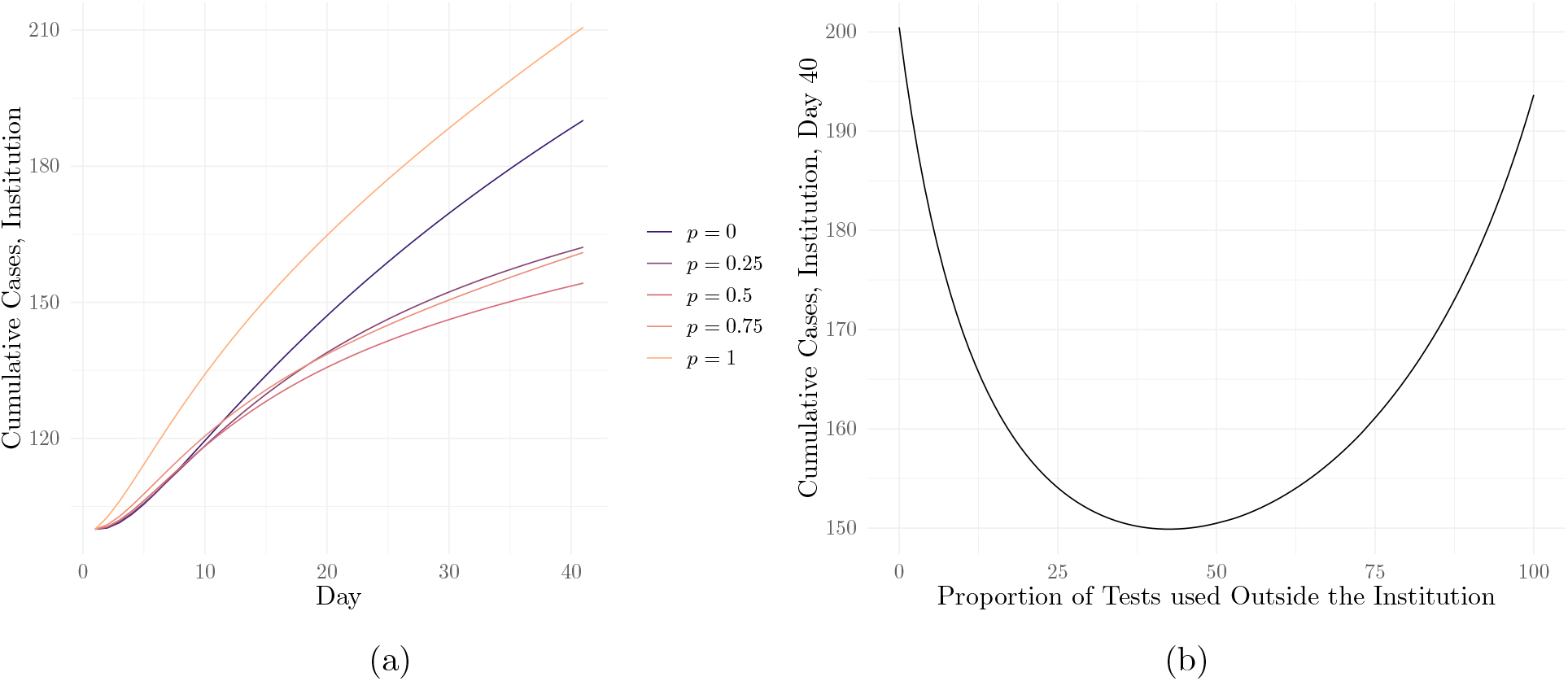
(a) Modeled cumulative cases over time at CMU under 5 different proportions *p* of peripheral testing; (b) cumulative cases on day 40 as a function of the proportion of tests deployed to the periphery, with the minimum at 45% peripheral testing

Because of the potential limitations and biases of our CMU data, and because our model relies on numerous parameters that vary widely between institutions, we proceeded by demonstrating robustness to and characterizing the influence of several factors on our results. These included four key factors: local community prevalence, social mitigation efforts, testing capacity, and contact tracing adoption (see Figure 3). Of course, there are more factors to assess, including distributions in numbers of contacts, variance in transmissibility, and initial prevalence within the institution. For an assessment of these factors, see Appendix B, or, for an interactive sensitivity analysis, visit https://ispecht.shinyapps.io/covid19-altruistic-testing/.

**Figure 3:**
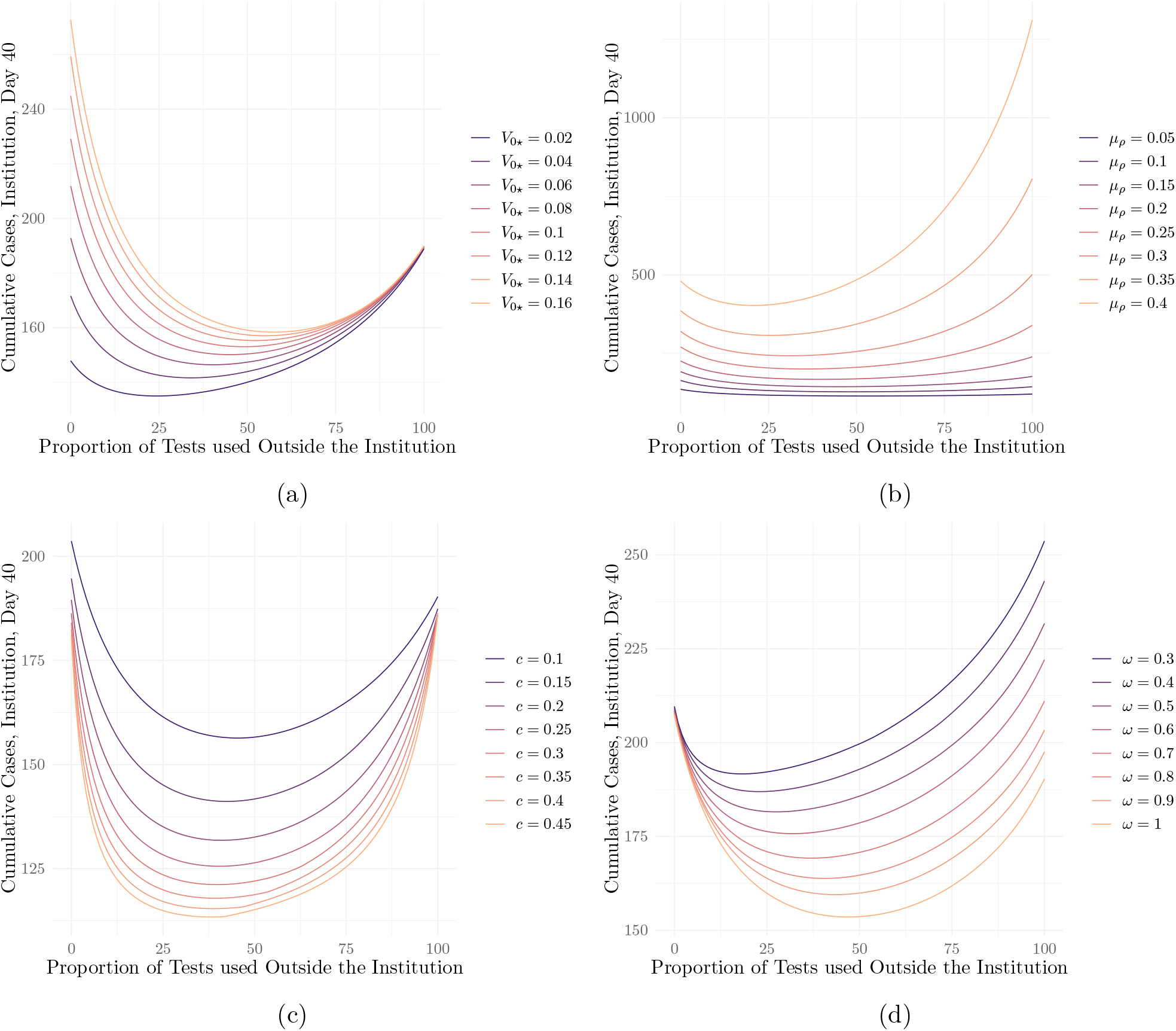
Cumulative cases on day 40 as a function of the proportion of tests deployed to the periphery under different values of (a) the initial prevalence in the periphery, *V*_0*\**_; (b) the secondary attack rate among institution members, *µ*_*ρ*_; (c) the tests-per-person-per-day ratio, *c*, and (d) the proportion of contacts traced *ω*

We first assessed sensitivity to the prevalence of COVID-19 among members of the periphery (*V*_0*\**_). While the 6% positivity rate in Mesa County in January 2020 was reflective of national statistics at the time, different institutions faced significantly higher or lower caseloads in their surrounding communities, representative of factors such as density and public health resources that varied from place to place. Beyond that, rates have naturally varied over the course of the outbreak. We observed that under higher values of *V*_0*\**_, the effectiveness of redistributing tests to the periphery grew, with the optimal proportion increasing from 24% when *V*_0*\**_ = 2% to 58% when *V*_0*\**_ = 16% (see Figure 3a). This finding is unsurprising, given that the probability of any individual test detecting a case in the periphery grew with higher values of *V*_0*\**_. In turn, the resulting quarantine (of both the peripheral member and their contact in the institution) minimized the probability of the virus breaching the institution.

Next, we turned to social mitigation efforts. While several model parameters capture these efforts, we focused on the secondary attack rate (SAR) for institution members––that is, the probability of transmission between an infectious individual and a susceptible one. Individuals may decrease the proportion of contacts they infect by, for example, wearing masks and socially distancing––thereby decreasing the observed SAR. Many institutions employ different mitigation measures, but altruism proved an effective strategy under a wide range of values for the observed SAR among institution members, denoted *µ*_*ρ*_ (see Figure 3b). For *µ*_*ρ*_ = 0.05, the optimal proportion of peripheral testing lay at 54%; this proportion decreased to 25% when *µ*_*ρ*_ = 0.4. This again fell in line with our expectations, as for higher *µ*_*ρ*_ values, a single case within the institution had much greater potential to spread, limiting the effectiveness of peripheral testing and lowering the optimal proportion of tests to be administered outside the institution. On the other hand, when *µ*_*ρ*_ was low, a single case within the institution likely did not give rise to an outbreak. This made institutional testing less essential, leaving more capacity to establish a ‘barrier of defense’ in the periphery to prevent cases from breaching the institution in the first place.

We then focused on *c*, the number of tests administered by the institution per day per person. An intuitive way to think about this parameter is that on average, an institution member receives a diagnostic test every 1/*c* days. We found that test redistribution to the periphery remained an effective strategy even for relatively low values of *c*, such as *c* = 10% (see Figure 3c). The optimal proportion of peripheral testing stayed relatively constant, ranging from about 38% to 48% for values of *c* between 45% to 10%. This result reflected the fact that under our baseline CMU parameters, institution members average about 10 times more contacts within the institution than outside it. As such, the size of the periphery was small, limiting the possible pathways for the virus to breach the institution. Our results tell us that even with limited testing resources, tests would best be used to prevent the virus from entering the institution via these pathways. Note that we also investigated different distributions of contacts within and outside the institution; for an analysis, see Appendix B, Figures S1a-S1d.

Finally, we accounted for the fact that institution members may not report all of their contacts in the periphery, or may have contacts that the institution cannot test due to factors such as geographic disparity. We captured the proportion of reported, testable contacts with the parameter *ω*. As our results suggest, contact reporting needed not be perfect for peripheral testing to help curb viral spread (see Figure 3d). Even if institution members reported only 30% of their contacts, the optimal proportion of peripheral testing lay at 18%; this proportion increases to 45% as the fraction of reported contacts grows to 100%. This result makes sense because the most socially-active––and therefore riskiest––members of the institution had many peripheral contacts, at least some of which will likely be known to the institution even under imperfect contact tracing (e.g. familial contacts). A positive test from even just one of these contacts would send the original institution member into quarantine, allowing the ‘barrier of defense’ strategy to remain an effective means of protecting the institution.

We note that the model exhibited some slight variance between runs due to the stochastic nature of the contact network generation step. While such stochasticity slightly affected numerical values for case counts between model runs, the shapes of the curves in Figures 2-3 remained consistent.

## 4 Discussion

Our model supports our hypothesis that the altruistic approach––in which institutions test beyond their walls––is the most effective protection strategy. In every instantiation of the model, we observed that deploying some proportion of diagnostic tests to the periphery significantly reduces the cumulative caseload at the end of a 40-day period. The optimal proportion of peripheral testing was 45% under baseline parameters and ranged from 18% to 58% under different levels of local prevalence, social interaction, testing, and contact tracing.

Our methods serve as a general framework for modeling one specific population within the context of another, and we hope that further research may help refine the intricacies of such dynamics. We also hope our work provides justification for institutions to consider implementing an altruistic testing strategy, and for legal and regulatory bodies to create a path for them to do so. We encourage institutions to partner with local public health authorities to support testing or connect members of the periphery with the appropriate testing provider, as Colorado Mesa University and the University of California Davis have done [4, 8].

Our results urge a fundamental rethinking of how institutions with substantial testing capacity approach safety amid outbreaks. Epidemics are one of those rare instances where a seemingly selfless approach is, in fact, the most self-serving: institutions must help test beyond their walls to stay safe within them.

## Data Availability

Data from CMU and code used to implement the model and generate Figure 2a is available at https://github.com/broadinstitute/covid19-altruistic-testing.

## Ethics Statement

The study received a “Not Human Subjects Research” determination by the Broad Institute’s Office of Research Subject Protection.

## Acknowledgements

We would like to acknowledge the following individuals for their support and feedback for this work: Kristian G. Andersen, Tim Foster, Gabrielle Gionet, William Hanage, Daniel Larremore, David O’Connor, Roy R. Parker, and Megan Vodzak, as well as members of the Sabeti Lab, the Broad Institute, Colorado Mesa University, COVIDCheck Colorado, and Fathom Information Design. This work was made possible by the Gordon and Betty Moore Foundation, Howard Hughes Medical Institute, Flu Lab, and a cohort of generous donors through TED’s Audacious Project, including the ELMA Foundation, MacKenzie Scott, the Skoll Foundation, and Open Philanthropy.

## A Methods

### Overview

Here we derive the full set of modeling equations we used to conduct our analysis. We construct an agent-based epidemiological model (ABM) with known contact network in order to compare the effectiveness of various testing regimes at controlling viral spread within an institutional context. Specifically, we model two groups of agents—the members of an “institution” (such as a school, business, etc.) and all of their close contacts outside the institution (henceforth referred to as the “periphery”)—and assume the institution has access to a disproportionately-large tests-per-person-per-day ratio in comparison to the periphery. We then analyze the effect of redistributing some of the institution’s testing capacity to the periphery. The model provides key insight into the optimal proportion of tests to be redistributed in such a way. More broadly, the set of equations we provide serve as a general framework by which institutions may assess the relative effectiveness of various testing protocols.

We simulate an institution with *N* agents that interact according to a random graph whose node degree distribution reflects known contact patterns of the institution’s members. Let *M*_*i*_ be a random variable representing the number of edges connected to node *i*. To reflect the overdispersion typically associated with node degrees in social contact networks, we assume that the *M*_*i*_’s are i.i.d. negative binomial with mean *µ* and variance *σ*^2^ [12]. We can express *M*_*i*_ as a function of the activity levels **a** = (*a*_1_, …, *a*_*N*_) of the *N* individuals—parameters that represent sociability, whose distribution we will derive. Assuming proportionate mixing, we define *E*_*ij*_, the probability of nodes *i* and *j* sharing an edge, to be

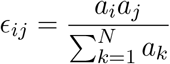

for *i* /= *j*, and 0 for *i* = *j*. Assuming that *E*_*ij*_ is small in general (an assumption we will justify below), we have that 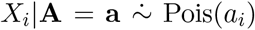, where 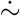 means “approximately distributed as.” So, we now need to choose a distribution for the *A*_*i*_’s to ensure that the marginal distribution of *X*_*i*_ is Negative Binomial. Since the Negative Binomial distribution may be expressed as the Poisson distribution compounded with the Gamma distribution, we model the *A*_*i*_’s as i.i.d. Gamma(*α, λ*) random variables. To solve for the parameters, we have that

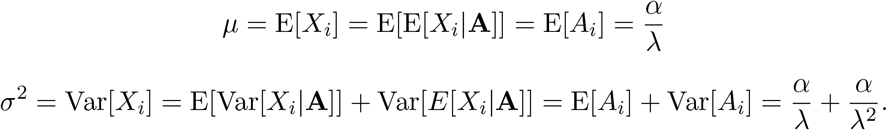

Thus,

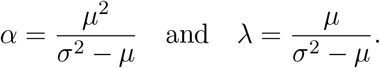

Finally, the assumption that 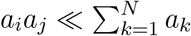 follows from the Cauchy-Schwarz inequality. We have that:

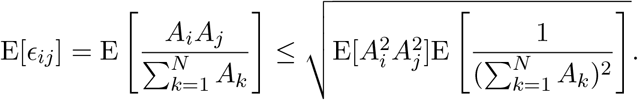

For the first expectation within the radical, we have that

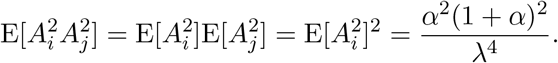

For the second expectation, let 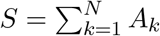. Then *S* ∼ Gamma(*Nα, λ*). Therefore

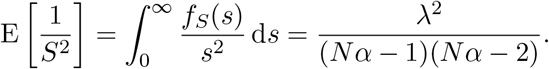

so long as *Nα >* 2. Thus,

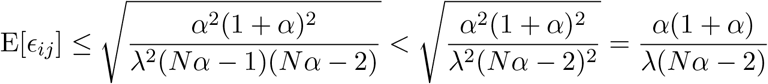

which approaches 0 as *N* → ∞.

Thus, we may generate our contact network by drawing the *a*_*i*_’s from the Gamma(*α, λ*) distribution, then drawing a Bern(*E*_*ij*_) random variable for each unordered pair of nodes {*i, j*} to generate the edges of the graph stochastically. Note that the probability of *E*_*ij*_ exceeding 1 is small because *P* (*E*_*ij*_ ≥ 1) ≤ E[*E*_*ij*_] by Markov’s inequality; in practice, this never occurred for all parameter combinations studied in this paper.

For contacts outside the insitution, we assume for simplicity that no two agents share a peripheral contact. Letting *Y*_*i*_ be the number of close contacts in the network of agent *i* outside the institution, we model the *Y*_*i*_’s as i.i.d. Negative Binomial random variables with mean *µ*_*\**_ and variance 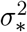. Contact reporting outside the institution, however, is likely imperfect—and even among reported contacts, some may be unwilling to receive a diagnostic test. To address this, we introduce the parameter *ω*, which represents the probability of any given contact outside the institution being traced and testable by the institution. Then, letting *Z*_*i*_ be the number of traced and testable contacts made by agent *i* outside the institution, we model *Z*_*i*_|*Y*_*i*_ ∼ Bin(*Y*_*i*_, *ω*). Moving forward, we will treat the contact network as fixed, and thus we will use the notations **x, y**, and **z** to represent the values taken on by random variables **X, Y**, and **Z**. We refer to the set of all traced and testable people outside the institution as the “known periphery”

### Viral propagation model

To capture the structure of a given graphical contact network, we take an agent-based approach for members of the institution; for the periphery, we implement a compartmental model that describes the population at large. Moreover, we assume that the prevalence in the periphery exhibits an epidemiological steady-state, a somewhat atypical assumption that we will explain further below and test as part of our sensitivity analysis (see Figure 3a and Appendix B). Finally, we include inter-compartmental “flux” terms in our modeling equations to account for the propagation of cases from the periphery into the institution.

We begin with the compartmental model for the periphery. We model five compartments: Susceptible (*S*_*\**_), Exposed (*E*_*\**_), Infectious (*V*_*\**_), Exposed/Infectious and Quarantined (*Q*_*\**_), and Recovered/Deceased (*R*_*\**_). Note that from now on, we will use the lower star notation for variables/parameters specific to the periphery. For our purposes we take exposed to mean “contracted the virus, but not yet infectious,” and assume that every agent enters the exposed stage before entering the infectious stage [6]. Let *γ* and *δ* be the recovery rate and exposed-to-infectious transition rates, respectively. Let *c* be the tests-per-person-per-day ratio, of which a proportion *p* are designated for members of the known periphery. Assuming for simplicity that each member of the known periphery is equally likely to be tested on any given day, we may set *η*_*\**_, the daily probability of an agent in the known periphery being tested, as follows:

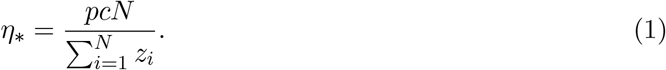

We only need to compute the steady-state probabilities of being exposed and infectious in the periphery, since the other states do not substantively impact our model. We assume that *prior to interventions*, i.e. testing and quarantine, the steady-state probability of being infectious is *V*_0*\**_. Given the transition rates, we may then assume that *E*_0*\**_, the steady-state probability of being exposed prior to interventions, is *γV*_0*\**_*/σ*. Finally, since *γV*_0*\**_ recover per day, it follows that *γV*_0*\**_ must transition from susceptible to infected each day to maintain the steady state.

Next, we include the effects of interventions, which affect only the known periphery. COVID-19 diagnostic tests have been shown to be highly reliable during the infectious stage of illness, but often fail to detect the extremely low viral loads present during the exposed staged [6]. As such, we let *ψ*_*E*_ be the test sensitivity during the exposed stage and *ψ*_*V*_ be the test sensitivity during the infectious stage. Modeling the exposed-to-quarantined and infectious-to-quarantined rates as *η*_*\**_*ψ*_*E*_ and *η*_*\**_*ψ*_*V*_, respectively, we arrive at the following system of differential equations:

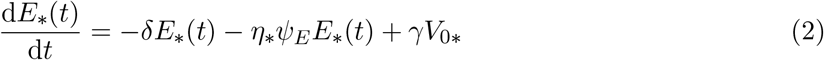

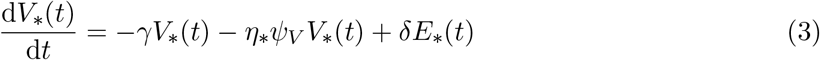

where *E*_*\**_(*t*) and *V*_*\**_(*t*) are the probabilities of an individual in the known periphery being exposed and infected at time *t*, respectively. Setting both derivatives equal to 0 and solving in terms of *V*_0*\**_, we obtain the steady states, which we denote *E*_*∞\**_ and *V*_*∞\**_, respectively:

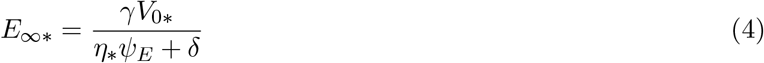

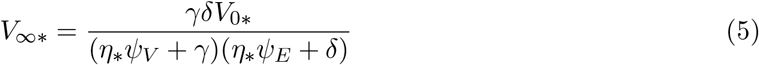

In the “unknown periphery” (i.e. members of the periphery who institution members do not report), we may assume that the the probabilities of being exposed and infected are *E*_0*\**_ and *V*_0*\**_, respectively.

We now turn our focus to the institution. Given the level of granularity provided by contact networks, along with the difficulty in capturing agent-specific conditions through more traditional epidemiological methods (i.e. compartmental modeling for the population at large), we model the state of each agent in the institution probabilistically using *N*-Intertwined Mean-Field Approximation (NIMFA) [17]. A widely-used and computationally-efficient approximation for the true stochastic process, the NIMFA approximates the joint probability of some agent *i* being susceptible and some other agent *j* being infectious as the product of the marginals. In its original form, the NIMFA states that:

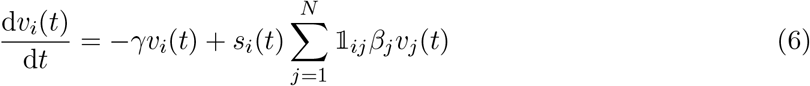

where *v*_*i*_(*t*) and *s*_*i*_(*t*) are the probabilities of agent *i* being infectious and susceptible at time *t*, respectively; 𝟙_*ij*_ is the indicator of an edge between *i* and *j*; *γ* is the recovery rate (as before), and *β*_*j*_ is the probability of transmission between infectious agent *j* and a susceptible agent per unit time.

We allow for heterogeneity in ***β*** = (*β*_1_, …, *β*_*M*_), where *M* is the total number of agents in both the institution and the periphery, to account for part of the overdispersion in COVID-19 transmission (the other part coming the node degree distribution in our graphical model). To derive ***β***, we begin by drawing ***ρ*** = (*ρ*_1_, …, *ρ*_*M*_), the (heterogeneous) secondary attack rate specific each agent, from a Beta distribution with mean *µ*_*ρ*_ and variance 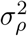. Since we defined *ρ*_*i*_ to be the probability of transmission over the course of agent *i*’s infection, and since the probability density of recovery at time *t* is equal to *γ* exp(−*γt*), we may write the total probability of transmission *ρ*_*i*_ in terms of the daily probability of transmission *β*_*i*_:

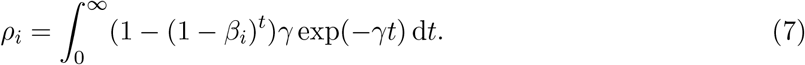

And, solving for *β*_*i*_, we obtain:

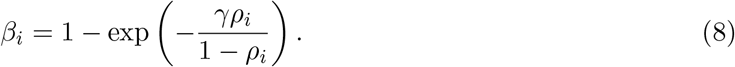

Note that in the case of COVID-19, limited literature exists on the variation in the secondary attack rate between agents, and the extent to which this phenomenon influences overdispersion is not known. As such, our distribution of the *ρ*_i_’s will be weakly informed, so we assess sensitivity to *µ*_*ρ*_ and 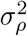 in Appendix B.

Having solved for ***β***, we now adapt the NIMFA to include additional transmission pathways and compartments for each individual agent while staying true to the original idea. Within the institution, we model seven compartments: Susceptible (*S*), Exposed (*E*), Infected (*V*), Recovered/Deceased (*R*), Susceptible and Quarantined (*U*), Exposed and Quarantined (*W*), and Infected and Quarantined (*Q*). See below for a diagram of all possible state transitions:

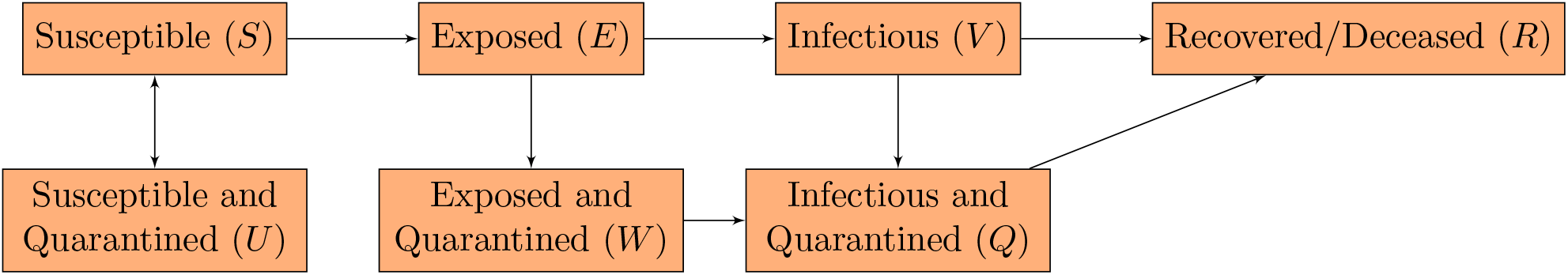

For any of the above states *Z* ∈ {*S, E, V, R, U, W, Q*}, we use the notation *z*_*i*_(*t*) to mean the probability that individual *i* is in state *Z* at time *t*. Since our agent-based model applies only to members of the institution, we allow *i* to range over integers only from 1 to *N*, inclusive. Revising (6) to account for the “exposed” compartment and for each agent’s outside-the-institution contacts, and then applying a Poisson approximation (to avoid the fact that the sum of the transmission probabilities may exceed 1), we obtain:

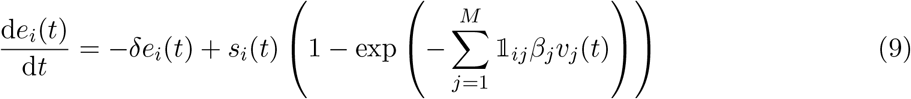

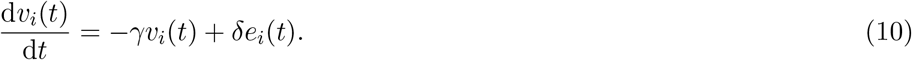

Note that for *j > N*, we take *v*_*j*_(*t*) to equal *V*_*∞\**_ if *j* is a member of the known periphery and *V*_0*\**_, otherwise. For ease of notation, from now on we let

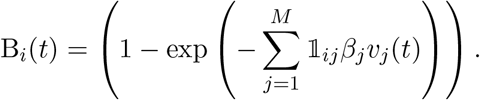

Next, we revise (9) and (10) to include the impact of quarantine due to a positive test result. To do so, we first need to decide on a prioritization for diagnostic tests within the institution. Let *η*_*i*_(*t*) be the probability that agent *i* is tested at time *t*. It is natural that we model this probability as roughly proportional to the number of known close contacts agent *i* has (both within and outside the institution) times the probability that agent *i* has not already been identified as a positive case. However, we also need to satisfy that the number of tests administered within the institution per unit time always sums to (1 − *p*)*cN*, and choosing a constant of proportionality to satisfy this condition may cause *η*_*i*_(*t*) to be greater than 1 for certain *i*. As such, we instead model *η*_*i*_(*t*) as

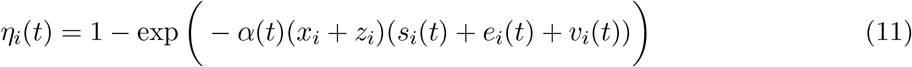

with *α*(*t*) chosen to satisfy

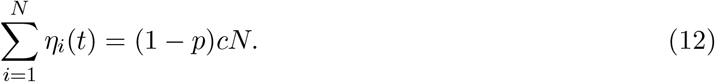

Note that no closed-form solution for *α*(*t*) exists; in practice, we compute it numerically for each discrete timestep Δ*t*. Just as in the model for the periphery, we then set the exposed-to-quarantined and infectious-to-quarantined rates (due to a positive test result) equal to *η*_*i*_(*t*)*ψ*_*E*_ and *η*_*i*_(*t*)*ψ*_*V*_, respectively. Now accounting for the quarantine rate due to a positive test result, we obtain:

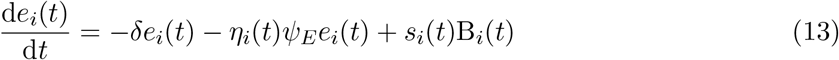

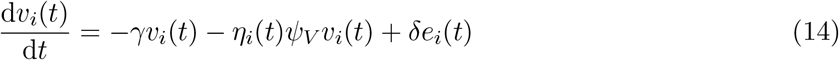

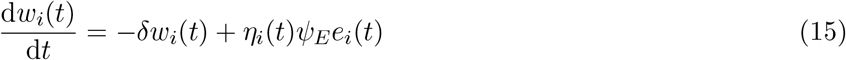

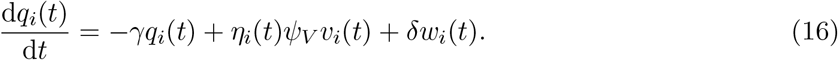

Finally, we include the effect of quarantine due to a positive test result from an agent’s first-degree contact. Again applying Poisson approximation to the events of each institutional agent’s first-degree contacts testing positive, we may write the time-dependent rate Γ_*i*_(*t*) at which agent *i* enters quarantine due to a known neighbor testing positive as:

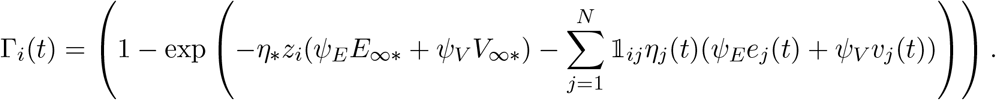

Revising (13-16) once again and adding in the remaining states, we arrive at our complete set of differential equations:

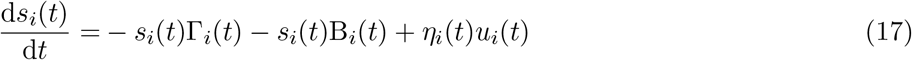

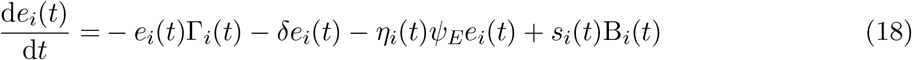

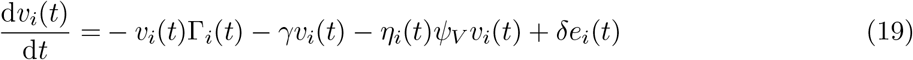

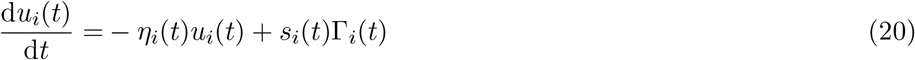

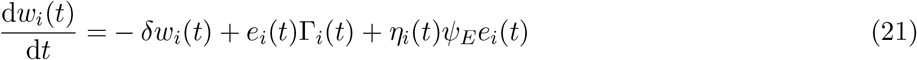

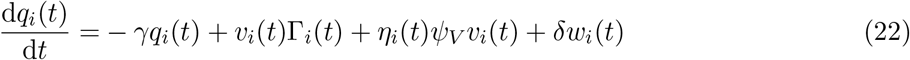

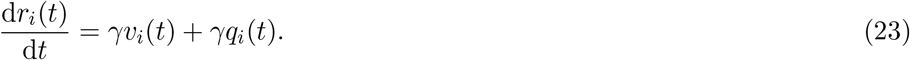

For the analyses conducted in this paper, we solved this system of differential equations using a discrete timestep of Δ*t* = 1 day.

## B Further Sensitivity Analysis

Here we build on the analysis in the Results section by assessing sensitivity to 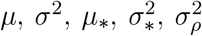, and **v**_0_, using a one-at-a-time approach. As in the Results section, for each parameter, we selected a range of possible values under which to plot cumulative cases on day 40 as a function of *p* (the proportion of tests deployed outside the institution). We found that the strategy of deploying at least some proportion of tests to the periphery is robust to a wide variety of parameter combinations (see Figure S1). Note that we observed some noise in our analyses of *µ*_*\**_ and 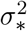 due to the skewness in the distribution of peripheral contacts; nonetheless, we still observed a clear trend in the day-40 cumulative case count as a function of peripheral testing under each values of *µ*_*\**_ and 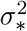 (see Figures S1c, S1d).

## C Peripheral Steady State

In our epidemiological model, we made the critical assumption that the periphery exhibits an epidemiological steady state. The rationale behind this assumption is that over the course of the COVID-19 pandemic, overall trends in cases (at the county, state, and national levels alike) have fluctuated based on a number of factors outside the control of individual institutions. As such, we chose not to model any change in the extra-institutional prevalence, but instead assumed it to be constant and then assess sensitivity to this factor (see Figure 3a). Here we provide additional analysis of how long after the implementation of testing in the periphery it takes in order to achieve such a steady state. Numerically approximating *E*_*\**_(*t*) and *V*_*\**_(*t*) based on equations (2-3), we obtained the curves shown in Figure S2, which provide estimates for the probabilities of being exposed and infectious in the periphery as a function of the duration of testing. We set all other relevant parameters to their baseline values (see Appendix D).

**Figure S1:**
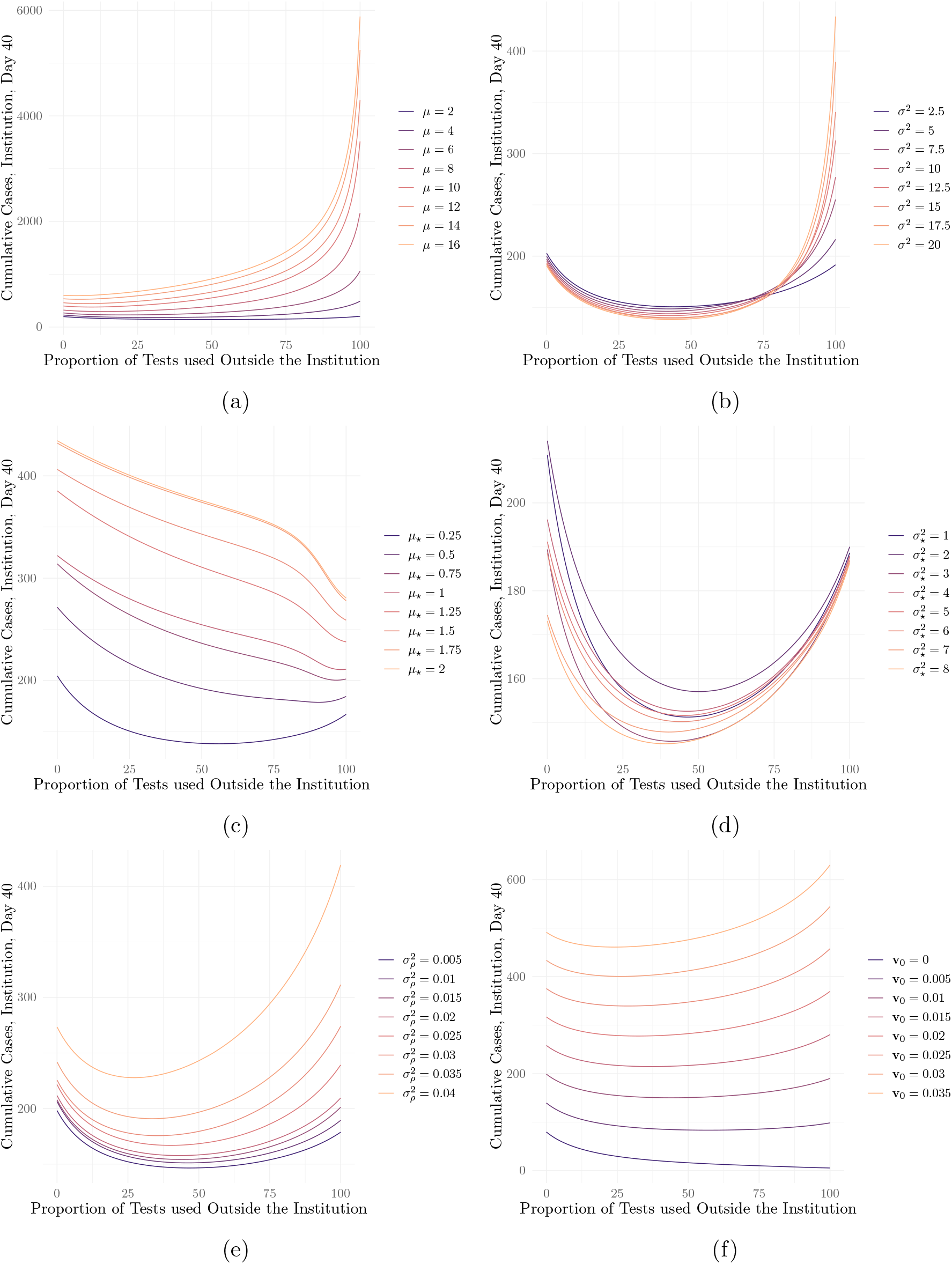
Cumulative cases on day 40 as a function of the proportion of tests deployed to the periphery under different values of (a) the mean number of institutional contacts, *µ*; (b) the variance in institutional contacts, *σ*^2^; (c) the mean number of peripheral contacts, *µ*_*\**_; (b) the variance in peripheral contacts, 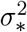; (e) the variance in the secondary attack rate, 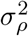; and (f) the initial prevalence at the institution, **v**_0_ (assumed to be uniform among agents)

**Figure S2:**
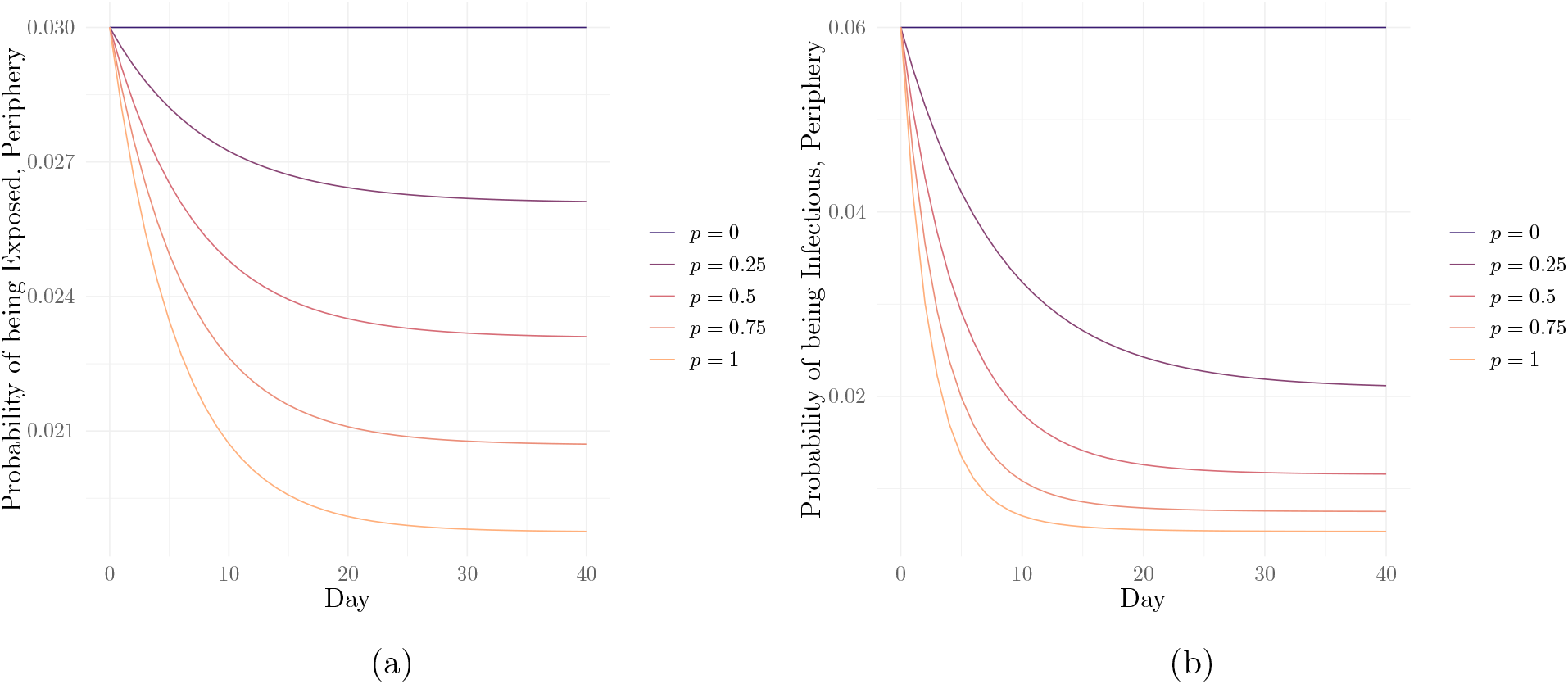
Modeled probabilities of an agent in the periphery being (a) exposed and (b) infectious over time starting with the implementation of community testing at time 0, under 5 different proportions *p* of peripheral testing.

## D Model Parameters

See below for a table of model parameters, explanations, values, and citations. Parameters with no citation are estimated based on data gathered at CMU. Parameters marked *\** are weakly-informed.

| Symbol | Description | Value | Citation |
| --- | --- | --- | --- |
| $N$ | Number of people in the institution | 10,000 | |
| $\mu$ | Mean number of contacts per agent at the institution | 2.3 | |
| $\sigma^2$ | Variance in number of contacts per agent at the institution | 2.4 | |
| $\mu_*$ | Mean number of contacts per agent outside the institution | 0.23 | |
| $\sigma_*^2$ | Variance in number of contacts per agent outside the institution | 1.8 | |
| $\omega$ | Proportion of contacts made outside the institution known to testing authorities | 1 | |
| $\mu_\rho$ | Mean secondary attack rate | 0.16 | [16] |
| $\sigma_\rho^2$ | Variance in secondary attack rate among agents | 0.01 | $\star$ |
| $\gamma$ | Recovery rate | 0.1 | [11] |
| $\delta$ | Exposed-to-infectious transition rate | 0.2 | [6] |
| $\mathbf{v}_0$ | Initial probability of infection within the institution, for each agent | 0.01 (uniform) | |
| $V_{0*}$ | Initial probability of infection outside the institution | 0.06 | |
| $c$ | Number of tests per day per person | 0.12 | |
| $\psi_E$ | Test sensitivity during the exposed stage | 0.02 | [10] |
| $\psi_V$ | Test sensitivity during the infectious stage | 1 | [10] |

